# Estimating the Transmission Potential of Symptomatic and Asymptomatic Cholera Cases from Household Microbiological and Clinical Data

**DOI:** 10.64898/2026.01.09.26343785

**Authors:** Claire P. Smith, Justin Lessler, Sonia T. Hegde, Taufiqur R. Bhuiyan, Taufiqul Islam, Faisal Ahmmed, Fahima Chowdhury, Ashraful I. Khan, Regina C. LaRocque, Richelle C. Charles, Ana A. Weil, Stephen B. Calderwood, Edward T. Ryan, Jason B. Harris, Andrew S Azman, Firdausi Qadri, Kirsten E. Wiens

## Abstract

**Background:** In Bangladesh, cholera treatment focuses on acute watery diarrhea in symptomatic cases at health facilities, though asymptomatic infections are common. Understanding the role of asymptomatic infections in transmission is crucial for designing appropriate control strategies in this setting.

**Methods:** We utilized data from household studies conducted in Dhaka, Bangladesh during 2006-2018 where a symptomatic confirmed cholera case and their household contacts were followed for thirty days. Vibriocidal antibodies, bacteriological culture, and symptom histories were collected at multiple times points. We used a hidden Markov model to estimate risk of infection from intra-household and extra-household (i.e., community and environmental) sources and to quantify relative risk of transmission from symptomatic and asymptomatic infected household contacts.

**Results:** Estimated daily risk of intra-household infection from a symptomatic individual to another household member was 2.6% (95% CI: 0.4% – 5.6%) and from an asymptomatic infected individual to another household member was 1.6% (95% CI: 0.2% – 4.5%). We found no significant differences in probability of infection from asymptomatic compared to symptomatic individuals (OR: 0.60; 95% CI: 0.11 – 3.23). We estimated that daily risk of infection from extra-household sources during follow-up was 1.0% (95% CI: 0.7% – 1.4%).

**Conclusion:** Mitigation measures focused solely on treatment of symptomatic cholera cases may be insufficient to prevent transmission in a household. This supports use of interventions that reduce risk of transmission irrespective of symptoms, such as prophylactic antibiotic treatment for household members and/or providing safe water and hygiene kits following a confirmed household or community case.

## Introduction

Seventh pandemic toxigenic O1 *Vibrio cholerae* is endemic in Bangladesh, which ranks as one of the highest cholera burden countries [1]. An estimated 66 million people are at risk and over 100,000 cases and 5,000 deaths occur annually [2]. These estimates, however, rely on clinical diagnosis of suspected cholera (i.e., acute watery diarrhea). A serological surveillance study in a cholera-endemic Bangladeshi community estimated that for every reported case, more than 2000 infections go unreported, and the vast majority of these are mild or asymptomatic [3]. Bangladesh is currently implementing a National Cholera Control Plan aiming for elimination by 2030 [4]. Since current treatment efforts focus on symptomatic cases that seek medical care, it is important to understand the role of asymptomatic infections in onward transmission.

Cholera control efforts can be further aided by understanding the relative contribution of intra- and extra-household transmission. Prior modeling work suggested that population vaccination could be more effective in reducing number of infections over a 40-day period compared to water treatment or antibiotic distribution when intra-household transmission is a significant epidemic driver [5]. Household contacts of cholera cases are at an increased risk of infection due to both proximal contact with infected household members (intra-household) and the potential for shared environmental and community exposures (extra-household) [1].

Household cohort studies estimated that risk of *V. cholerae* transmission from intra-household exposure to an infected household member is greater than risk of infection from sources outside the home [6], and genetic studies have found substantial levels of intra-household transmission [7,8]. However, these studies did not directly assess differences in transmission by symptom status.

Estimates of the proportion of *V. cholerae* infections that are symptomatic vary widely. Estimates from household cohorts and outbreak settings tend to be higher, ranging from 10% to 60% [9], while population-level serological surveillance and modeling studies imply that the true proportion of infections that involve symptoms may be less than 0.1% [3,10]. Mild and asymptomatic individuals often continue daily activities without antibiotic treatment, potentially increasing transmission opportunities, and the risk of infection from exposure to an asymptomatic contact remains unknown.

Studies of the duration of bacterial shedding, which is likely to be associated with increased risk of household transmission, yield inconsistent results; one, which followed household contacts of cholera cases over the course of ten days, found no significant differences in shedding by symptom status [11], while another found a notable difference in the number of positive cultures based on symptom status [12]. Stool volume and consistency also vary substantially between symptomatic and asymptomatic cases, which could influence transmission risk. Patients with acute watery diarrhea excrete high-volume stool, posing substantial risk for household surface contamination. Individuals with mild symptoms or asymptomatic infections produce lower-volume, more formed stools, potentially reducing opportunities for contamination and transmission. However, we know of no study that has attempted to directly measure differences in transmissibility between these groups.

Here we utilize data from two household-based studies in Dhaka, Bangladesh to estimate the risk of infection from intra- and extra-household sources and to quantify the relative risk of transmission from symptomatic and asymptomatic infected household contacts.

## Methods

### Study Design

We re-analyzed data from two household-based studies conducted in Bangladesh from December 2006 to January 2012, and February 2012 to September 2018 [11,13–15]. Culture-confirmed symptomatic index cholera cases were recruited when presenting for care at hospitals in Dhaka, Bangladesh. Consenting household contacts of the index case, defined as those with whom the index case shared a cooking pot for the three days prior to their enrollment, were enrolled on the following day. The index case and enrolled household contacts were followed for 30 days to assess infection and symptom status.

Three measurements of participants’ infection status over the follow-up period were analyzed in this study: presence of *V. cholerae* in culture of a stool or rectal swab, vibriocidal antibody titers, and self-reported symptoms. Rectal swabs for culture were taken on the first day of follow-up for index cases (i.e., the day the index case presented for care) and daily on days 2-10 of follow-up for household contacts (Figures 1, S1). Blood samples for immunological assays including vibriocidal antibody testing were taken on days 2, 7, and 30 for all participants. Presence of symptoms (watery diarrhea) was self-reported on days 1-10 and on day 30 for index cases and days 2-10 and day 30 for household contacts.

**Figure 1:**
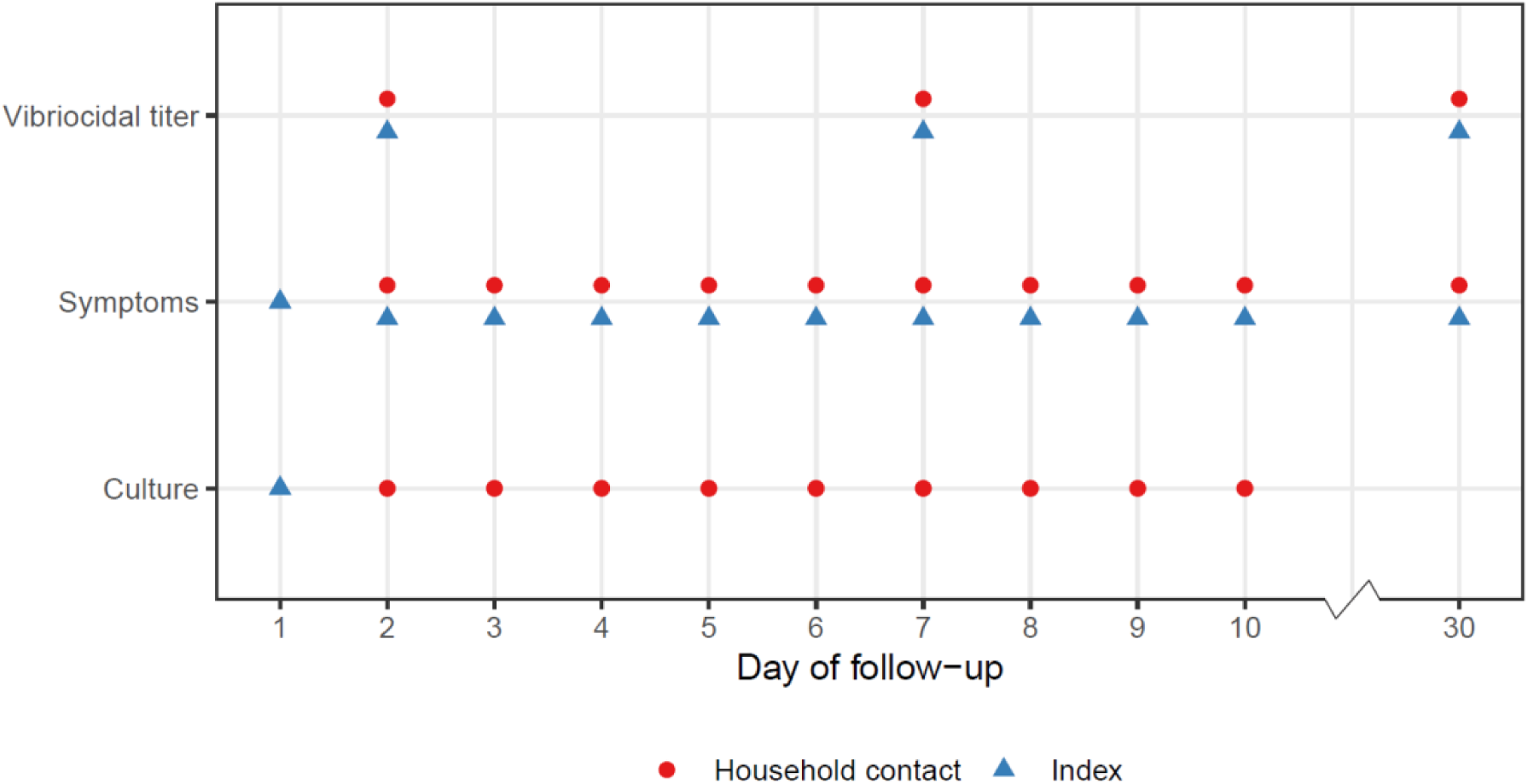
Sampling schedule for index cases and household contacts. Day 1 is the day that the index case presented for care.

At baseline, surveys were administered to collect individual- and household-level information, as previously described [11,13–15]. This analysis focuses on a subset of variables known or suspected to be associated with *V. cholerae* infection risk, namely age, sex, occupation, family and household structure, income, and access to water and soap [6,16–19]. Age group was categorized as 4 years of age and under, 5-17 years of age, and 18 years of age and older.

Occupation group was categorized as adults who work outside the home, adults who work at home, and children under 18 years of age. Familial relationship to the index case was categorized as self (for the index), spouse, parent, child, sibling, or unknown. We identified unenrolled household members as the difference between reported household size and the number enrolled in follow-up; these individuals were categorized as unknown for all demographic factors.

At the household level, the primary source of drinking water was recorded as boiled water, private tap, public tap, tubewell, or other. Monthly household income was recorded to the nearest 500 Bangladeshi taka (BDT). Household size and whether soap was used in the home was also recorded.

Laboratory procedures have been previously described [13,14,20]. Briefly, stool specimen/rectal swabs were cultured on taurocholate-tellurite-gelatin agar plates with confirmation of *V. cholerae* via slide agglutination. Vibriocidal antibody responses were measured using serial two-fold dilution; titers could take values ranging from 5 to 40,960. In our analysis, we used vibriocidal antibody responses for each household contact specific to the serogroup and serotype (O1 Inaba, O1 Ogawa, or O139) infecting the index case, as this was the serotype they were most likely exposed to within the household.

Approvals for the original studies were obtained from the institutional review boards of Massachusetts General Hospital and the International Centre for Diarrhoeal Disease Research, Bangladesh (icddr,b). Approval for re-analysis of the data in this study was obtained from Temple University.

### Statistical Analysis

We used a hidden Markov model [21–23] to estimate extra-household and intra-household infection risk by symptom status, factors modifying this risk, the proportion of infections that develop symptoms, and the duration of infectiousness. We estimate the intra-household infection risk based on the increased probability of infection in susceptible individuals exposed to another individual in their household actively infectious with cholera. Presumably such infections are due to transmission via shared food, water, and surfaces contaminated by the infectious household member. We attribute all other risk of infection to extra-household sources, which may include contaminated environmental sources in the community, community sources brought into the home (e.g., a contaminated water jug), water or food consumed outside the home, or exposure to infected community members through contaminated surfaces, facilities, or materials.

In this model, individuals can take on any of five latent states: susceptible (S), exposed but not infectious (E), asymptomatically infected (Ia), symptomatically infected (I_s_), and recovered (R), and transition between these states is illustrated in Figure 2. When an observation was available (reporting of symptoms, culture result, and/or vibriocidal titer), that individual’s probability of being in each of the latent states was reweighted according to the probability of seeing the observation if the individual is in that state. For example, the probability an individual is culture positive given they are in an infectious state is equal to the sensitivity of culture. This model was fit to individual-level data on report of watery diarrhea, culture results, and vibriocidal titer level and parameter estimates were obtained using the forward algorithm in the Stan probabilistic programming language [24].

**Figure 2:**
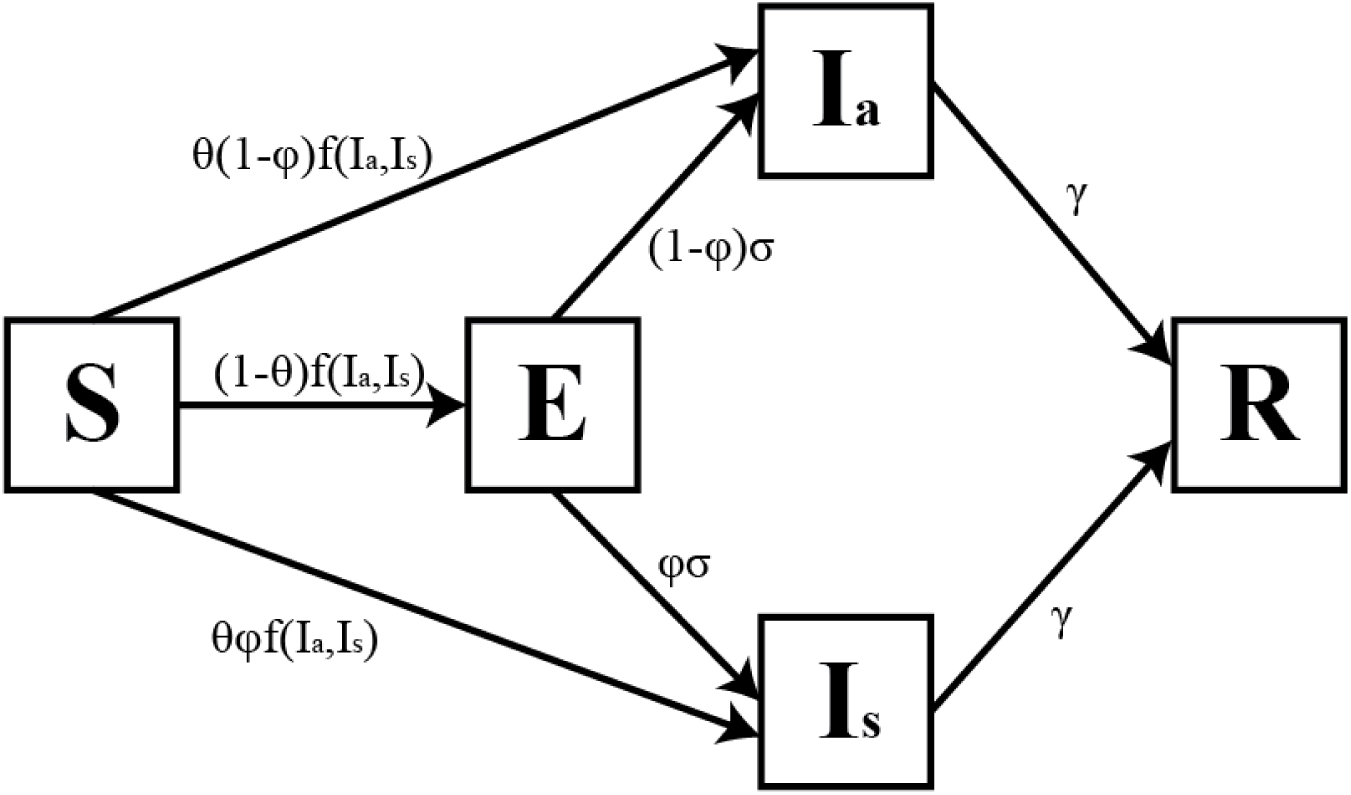
Infection process model for household cholera transmission. The function *f(I_a_, I_s_)* gives the probability of an individual becoming infected from an intra- or extra-household source (see Equation 2 in Supplemental Methods). φ is the probability of an individual becoming symptomatic given infection. θ is the proportion of cases with an incubation period less that one day. The rate at which exposed individuals become infectious is σ and the rate of recovery from infection is γ.

Intra- and extra-household infection risks were modeled as a logistic function of all individual- and household-level covariates. We assumed no interaction between symptomatic status and other covariates in their influence on intra-household transmission risk (i.e., symptom status was modeled as an independent risk factor). Full details of the model structure can be found in the Supplemental Methods.

At time points where an observation was available, state probabilities were updated based on the given observation. When individuals were in the S and E compartment, we assumed they had a 13% probability of symptoms (based on the proportion of household contacts with neither a positive culture results or four-fold rise in vibriocidal titer who reported symptoms during the follow-up period), nearly 100% probability of a low vibriocidal antibody titer, and a 5.7% probability of a positive culture result based on the specificity of culture [25]. When individuals were in either of the infected states, we assumed that the probability of a positive culture result was equal to the sensitivity of culture (82%) [25] and there was a 10% probability of a high titer response (based on prior descriptions of vibriocidal titer profiles in the days following infection [26]). Individuals in I_s_ were assumed to have a nearly 100% probability of reporting symptoms while individuals in I_a_ had nearly 0% probability of reporting symptoms. Individuals in R were assumed to have a 13% probability of symptoms, 70% probability of a high vibriocidal titer measurement, and a 5.7% probability of a positive culture result based on the specificity of culture [25]. Participants were classified as having a high titer response if the vibriocidal titer from the serum sample collected on the given day was greater than or equal to 320 [26]. Sensitivity analyses were performed to assess the impact of varying titer cutoffs.

We ran the following models: a baseline intercept-only model, univariate models for potential factors modifying transmission risk (age group, sex, occupation group, relation to the index case, monthly household income, drinking water source, and whether there was access to soap in the home), and a model adjusted for all factors except age. Age group was not included in the adjusted model due to collinearity with occupation group and familial relationship with the index case.

Models were validated using ten independently simulated data sets of 500 households each (see Supplemental Methods). Models were fit using Markov-chain Monte Carlo (MCMC) methods, with four chains each with 2000 iterations, of which the first half are discarded for warmup. We assessed convergence using visualization of trace plots and the Rubin-Gelman convergence statistic [27]. Point estimates are the median and credible intervals are the 2.5% and 97.5% quantiles of the posterior distribution. All models were run in stan (via Rstan version 2.32.7) [24] and all other analyses were conducted in R version 4.4.0 [28].

### Code and data availability

All analytical code and de-identified data are available at github.com/UNCIDD/vc-household.

## Results

There were 481 index cases and 784 household contacts enrolled across the cohorts over the analysis period. Households ranged from size 2 to 14, with the majority (54.3%) of households composed of 2-4 members (Table 1). The majority (60.1%) of households earned less than BDT 15,000 (approximately USD 123) in monthly income. The most common source of drinking water was a private tap (43.2%). The vast majority (95.4%) of households had access to soap in the home. All index and household infections were *V. cholerae* O1 serogroup.

**Table 1:** Household characteristics.

|  | **<u>Household</u>**<br>**<u>Characteristics</u>** |
| --- | --- |
|  | n = 481 |
| **<u>Household size</u>** |  |
| 2-4 | 261 (54.3%) |
| 5-7 | 190 (39.5%) |
| 8-10 | 27 (5.6%) |
| 11-14 | 3 (0.6%) |
| **<u>Monthly income (BDT)</u>** |  |
| 0-4,999 | 22 (4.6%) |
| 5,000-9,999 | 130 (27.0%) |
| 10,000-14,999 | 137 (28.5%) |
| 15,000-19,999 | 79 (16.4%) |
| 20,000+ | 113 (23.5%) |
| **<u>Drinking water source</u>** |  |
| Boiled water | 97 (20.2%) |
| Private tap | 208 (43.2%) |
| Public tap | 18 (3.7%) |
| Tubewell | 143 (29.7%) |
| Other | 15 (3.1%) |
| Use soap in home | 459 (95.4%) |

Of the 721 household contacts with an available culture result, 22.3% (161) had a positive culture result during the first 10 days of follow up (Table 2, Figure S2). Of these, 41.0% (66) reported experiencing watery diarrhea during this time period (Table 2). Of the 560 household contacts without a positive culture result (i.e., all culture results were negative), 12.5% (70) reported experiencing watery diarrhea during the first ten days of follow-up. Compared to those with all negative culture results, culture-positive household contacts had soap in the home less often (89.4% vs. 95.5%) and were less often in a household that used boiled water as their source of drinking water (11.2% vs. 20.5%). Culture-positive household contacts were more likely to be from lower-income households, with 52.2% residing in households earning less than BDT 10,000 per month compared to 35.0% for those without a positive culture.

**Table 2:** Enrolled participant characteristics stratified by participant type and culture result.

|  | All participants | Index cases | Household contacts |  |  |
| --- | --- | --- | --- | --- | --- |
|  |  |  | Culture + | Culture – | No culture |
|  | n = 1,265 | n = 481 | n = 161 | n = 560 | n = 63 |
| Mean age (IQR) | 26.3 (13.9-35.9) | 23.9 (10.4-33.2) | 24.9 (9.4-36.3) | 28.1 (18.8-36.4) | 31.3 (22.6-37.2) |
| <u>Sex</u> |  |  |  |  |  |
| Male | 669 (52.9%) | 293 (60.9%) | 74 (46.0%) | 270 (48.2%) | 32 (50.8%) |
| Female | 596 (47.1%) | 188 (39.1%) | 87 (54.0%) | 290 (51.8%) | 31 (49.2%) |
| <u>Occupation Group</u> |  |  |  |  |  |
| At home | 311 (24.6%) | 93 (19.3%) | 43 (26.7%) | 155 (27.7%) | 20 (31.7%) |
| Outside home | 599 (47.4%) | 228 (47.4%) | 58 (36.0%) | 274 (48.9%) | 39 (61.9%) |
| Under 18 | 355 (28.1%) | 160 (33.3%) | 60 (37.3%) | 131 (23.4%) | 4 (6.3%) |
| <u>Relation to index</u> |  |  |  |  |  |
| Parent |  |  | 56 (34.8%) | 220 (39.3%) | 27 (42.9%) |
| Spouse |  |  | 37 (23.0%) | 153 (27.3%) | 18 (28.6%) |
| Child |  |  | 35 (21.7%) | 91 (16.3%) | 7 (11.1%) |
| Sibling |  |  | 31 (19.3%) | 80 (14.3%) | 8 (12.7%) |
| Unknown |  |  | 2 (1.2%) | 16 (2.9%) | 3 (4.8%) |
| <u>Monthly household income (BDT)</u> |  |  |  |  |  |
| 0-4,999 | 62 (4.9%) | 22 (4.6%) | 9 (5.6%) | 30 (5.4%) | 1 (1.6%) |
| 5,000-9,999 | 375 (29.6%) | 130 (27.0%) | 75 (46.6%) | 166 (29.6%) | 4 (6.3%) |
| 10,000-14,999 | 361 (28.5%) | 137 (28.5%) | 41 (25.5%) | 173 (30.9%) | 10 (15.9%) |
| 15,000-19,999 | 190 (15.0%) | 79 (16.4%) | 16 (9.9%) | 75 (13.4%) | 20 (31.7%) |
| 20,000+ | 277 (21.9%) | 113 (23.5%) | 20 (12.4%) | 116 (20.7%) | 28 (44.4%) |
| <u>Drinking water source</u> |  |  |  |  |  |
| Boiled water | 245 (19.4%) | 97 (20.2%) | 18 (11.2%) | 115 (20.5%) | 15 (23.8%) |
| Private tap | 555 (43.9%) | 208 (43.2%) | 77 (47.8%) | 242 (43.2%) | 28 (44.4%) |
| Public tap | 42 (3.3%) | 18 (3.7%) | 5 (3.1%) | 19 (3.4%) | 0 (0.0%) |
| Tubewell | 388 (30.7%) | 143 (29.7%) | 55 (34.2%) | 170 (30.4%) | 20 (31.7%) |
| Other | 35 (2.8%) | 15 (3.1%) | 6 (3.7%) | 14 (2.5%) | 0 (0.0%) |
| <u>Soap in home</u> |  |  |  |  |  |
| Yes | 1,200 (94.9%) | 459 (95.4%) | 144 (89.4%) | 535 (95.5%) | 62 (98.4%) |
| No | 65 (5.1%) | 22 (4.6%) | 17 (10.6%) | 25 (4.5%) | 1 (1.6%) |
| <u>Symptoms during first 10 days</u> |  |  |  |  |  |
| Watery diarrhea only | 215 (17.0%) | 121 (25.2%) | 41 (25.5%) | 49 (8.8%) | 4 (6.3%) |
| Vomiting only | 38 (3.0%) | 17 (3.5%) | 3 (1.9%) | 18 (3.2%) | 0 (0.0%) |
| Watery diarrhea & vomiting | 100 (7.9%) | 50 (10.4%) | 25 (15.5%) | 21 (3.8%) | 4 (6.3%) |
| None | 903 (71.4%) | 287 (59.7%) | 92 (57.1%) | 470 (83.9%) | 54 (85.8%) |
| Missing | 9 (0.7%) | 6 (1.2%) | 0 (0.0%) | 2 (0.4%) | 1 (1.6%) |

In the baseline model, the estimated per-day probability of infection from extra-household sources was 1.0% (95% credible interval [CI]: 0.7% – 1.4%). The estimated per day risk of intra-household infection was 2.6% (95% CI: 0.4%-5.6%) if the infectious household member was symptomatic and 1.6% (95% CI: 0.2% – 4.5%) if they were asymptomatic (Figure 3). We estimated that the probability of an infection resulting in development of symptoms was 34.0% (95% CI: 20.4% – 53.0%) and the average duration of infectiousness was 2.2 days (95%CI: 2.1 – 2.4).

**Figure 3:**
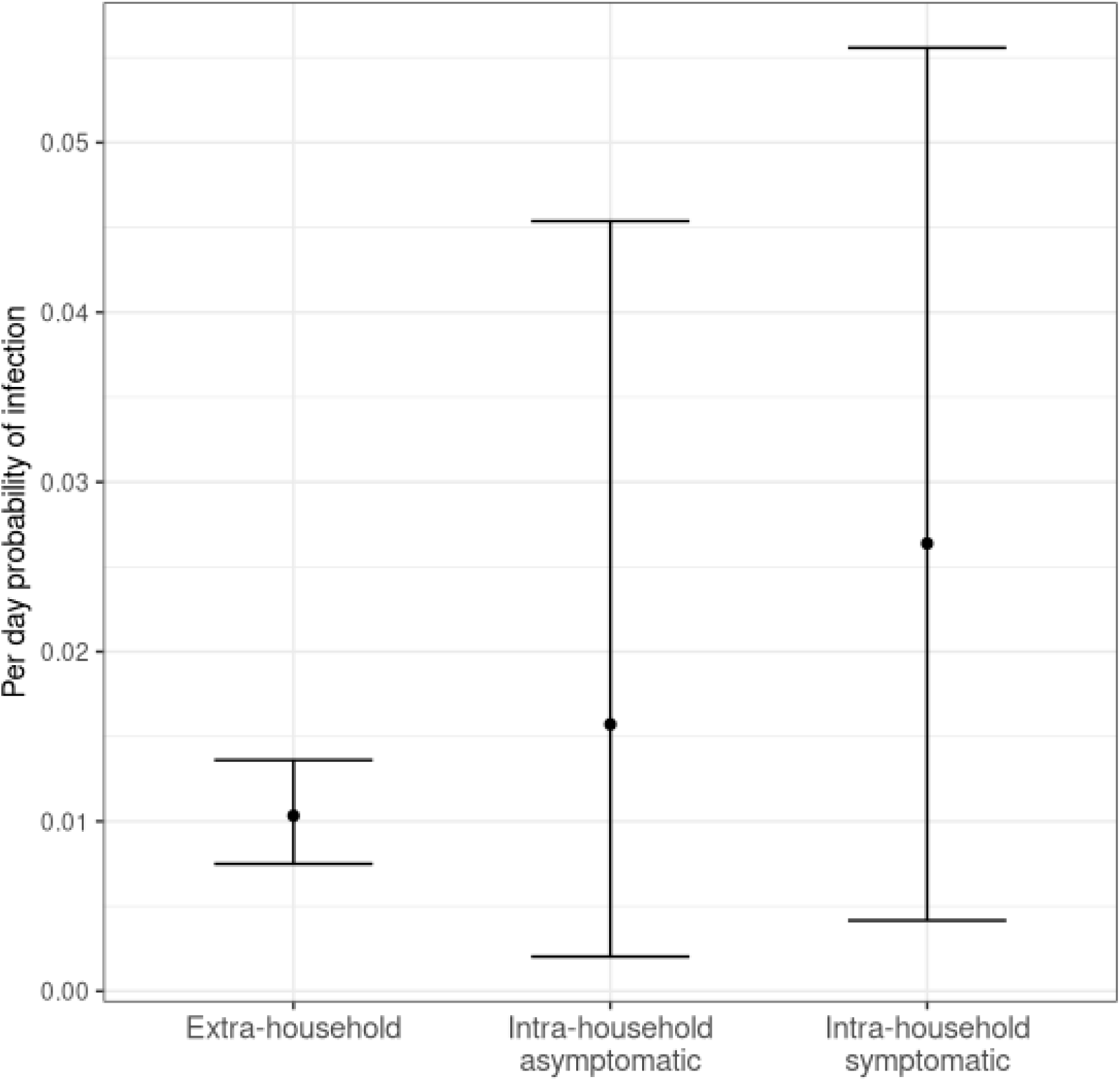
Daily probability that a susceptible individual is infected by a source other than an infectious household member (“extra-household”), an asymptomatically infected household member (“intra-household asymptomatic”), and a symptomatically infected household member (“intra-household symptomatic”).

We found no significant difference in the probability of infection following exposure to an asymptomatic versus symptomatic household member. In the baseline model, the daily odds of infection from an asymptomatic household member was 0.60 (95% CI: 0.11 – 3.23) times the daily odds of infection from a symptomatic household member. Based on the posterior distribution of baseline model parameters, there is a 73.7% chance that the probability of infection from a symptomatic household member is greater than that of an asymptomatic household member (Figure S3). The relationship remained not statistically significant after adjusting for participant and household characteristics (odds ratio [OR] 0.61, 95% CI 0.10 – 3.98).

In adjusted analyses, we found several factors associated with extra-household infection risk. Compared to those who worked outside the home, extra-household infection risk was elevated for adults who worked in the home (OR: 2.33, 95% CI: 1.13 – 5.31) and for household members under age eighteen (OR: 1.50, 95% CI: 0.57 – 4.12) (Figure 4, Table S1). Those who lacked access to soap in the home had 2.03 (95% CI: 0.89 – 4.41) times the daily odds of acquiring an infection from an extra-household source.

**Figure 4:**
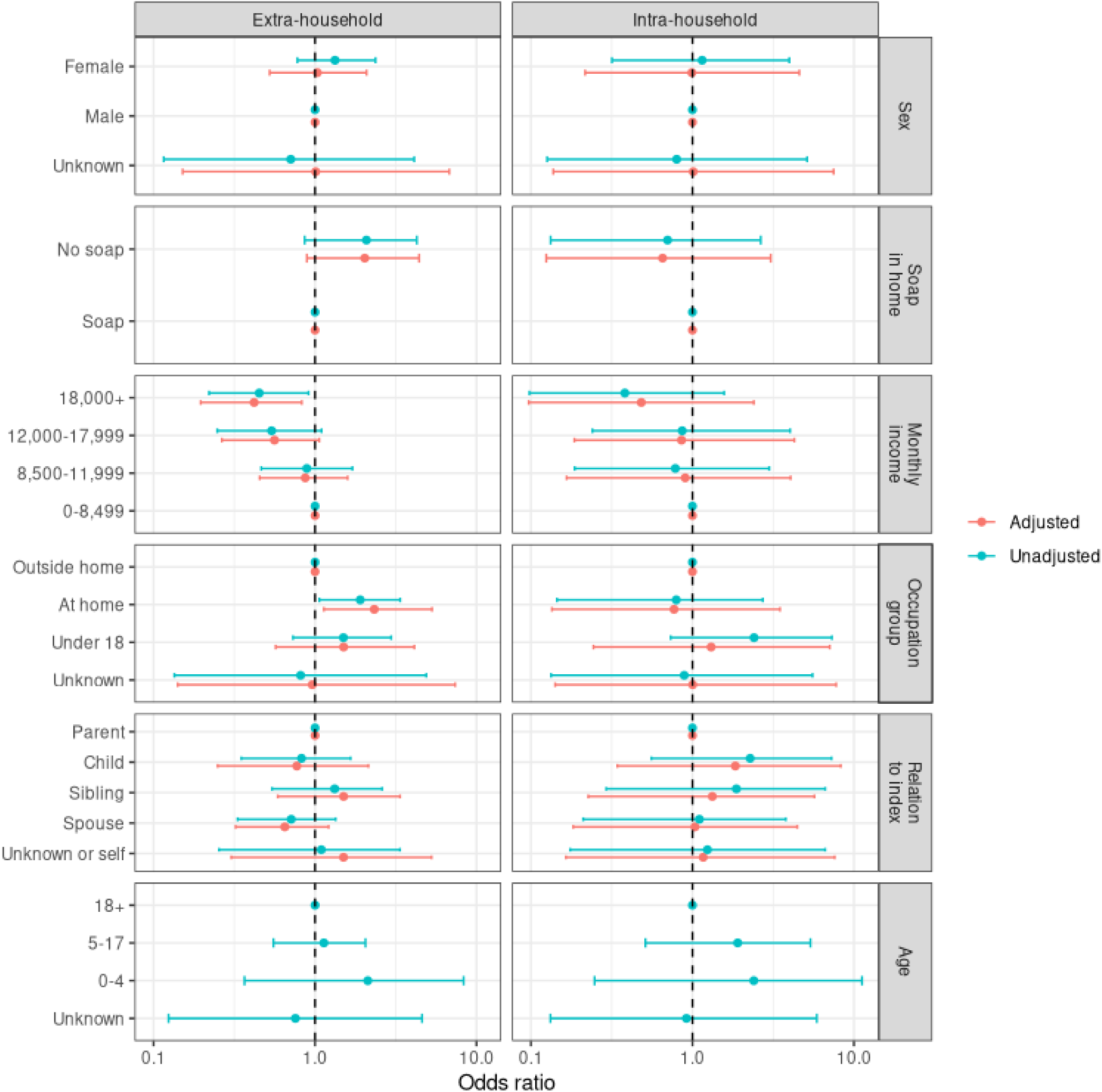
The relative odds of daily intra- and extra-household infection by sex, drinking water source, use of soap in the home, monthly household income (BDT), occupation group, relation to index, and age group. The adjusted model includes all covariates except age group. Shown are medians and 95% credible intervals. Reference categories are denoted by a point estimate at 21.0 without a credible interval. Socio-demographic data is missing (“unknown”) for unenrolled household members, as described in the methods.

Compared to households earning less than BDT 8,500 per month, households who earned BDT 18,000 or more (the highest income quartile) had 58% lower daily odds (OR: 0.42; 95% CI: 0.20 – 0.83) of extra-household infection. Although not statistically significant, individuals in households who earned BDT 18,000 or more per month also had the lowest risk of intra-household infection (OR: 0.48; 95% CI: 0.10 – 2.40 compared to those earning less than BDT 8,500 per month) (Figure 4, Table S1).

In sensitivity analyses, raising the cutoff for a vibriocidal titer response indicative of recovery (i.e., recent infection) decreased the daily extra-household infection risk but did not significantly alter daily intra-household infection risks, the symptomatic proportion, nor the duration of infectiousness (Figure S4). Both increasing the probability that the index case started in the symptomatically infected compartment and increasing the probability of observing symptoms in uninfected individuals decreased the symptomatic proportion but did not significantly alter other parameter values (Figure S5-6).

Validation experiments showed that our model was able to successfully recover the underlying parameters in simulated data. For 100% of the simulated datasets, the 95% credible intervals for the infection probability parameters contained the true value (Figure S7). The model was also able to accurately recover the number of individuals in each latent state (Figure S8).

## Discussion

We found no significant differences in daily infection risk from symptomatic and asymptomatic infected household members. This implies that a passive system of symptom-based surveillance may fail to capture many infections that are actively contributing to transmission. Testing and treatment of high-risk contacts, including asymptomatic individuals, may therefore complement prevention strategies such as vaccination and improved access to clean water and hygiene resources. Furthermore, while our findings confirm the acute risk of cholera infection posed by household members, they also highlight the high risk of infection from other sources likely experienced by members of households where cholera cases are present.

The period when a cholera index case is identified in a household will be, in most cases, a period when other household members are at elevated risk of getting cholera outside of their household. This is reflected in our results, which estimate a 26% chance of extra-household cholera infection for the index case’s household members during the subsequent month.

However, when a household member is shedding live *V. cholerae*, they remain the most likely source of infection and their presence doubles the risk of infection in household members on any given day during their infectious period (more so if multiple infectious individuals are present).

Our finding that those who work at home are at elevated risk of extra-household infection may at first seem counterintuitive. However, it is important to remember that this risk captures the chances of infection from all sources other than an infectious household member, and those who work at home may have more frequent exposure to shared environmental and community sources near the home. For instance, the household’s drinking water may have been contaminated from an extra-household source, and prior studies have found that the presence of *V. cholerae* in a household’s water source and stored drinking water (from any source) was associated with significantly higher odds of *V. cholerae* infection [16,29].

Low socioeconomic status is a known risk factor for *V. cholerae* infection [17–19] and we observed decreased extra-household infection risk with increased monthly income, suggesting that increasing socioeconomic status is associated with protection against infection from extra-household sources. Higher socioeconomic status may also be associated with reduction in the risk of intra-household transmission. This protective effect may be due to more consistent access to clean water and sanitation supplies and reduced likelihood of living in highly crowded conditions that facilitate transmission [19,30].

This study is subject to several limitations. Information about index cases was partially censored because they were enrolled partway into their infection period, and we lacked culture results after enrollment. All index cases received antibiotics before returning home, which significantly reduces bacterial shedding and may affect infectiousness [31,32]. Additionally, the sensitivity of vibriocidal antibody titers to detect asymptomatic infections is unknown in this population. We attempted to address some of these limitations in the design of the model; for example, by modeling the day before index enrollment to account for uncertainty regarding the duration of index case symptoms when they presented for care. Our confidence in the results is further supported by our estimates of key natural history parameters such as duration of infectiousness (2.2 days) and the symptomatic proportion (34%) are broadly consistent with previous estimates [9,11].

We assumed that symptomatic and asymptomatic cases recovered at the same rate, although data on duration of infection by symptom status is limited and this assumption may not be valid. In addition, we did not account for seasonal or other temporal fluctuations in extra-household infection risk, which can be substantial in this setting [33,34]. Future extensions of this model may use background case counts or seasonal terms to inform a time-varying extra-household infection probability.

Our results underscore the important role that both symptomatic and asymptomatic cases play in *V. cholerae* transmission and suggest that environmental persistence of *V. cholerae* may heighten infection risk in homes with recent infections. This suggests that effective cholera surveillance and control efforts should target both asymptomatic and symptomatic cases while extending prevention efforts beyond the resolution of primary infections. Strategies such as highly targeted prophylactic antibiotic treatment during high-risk periods, provision of safe water options (including chlorine tablet distribution), and hygiene kit distribution may reduce cholera burden in Bangladesh and other endemic settings. Because undetected asymptomatic cases pose a risk of onward transmission, these efforts should, where feasible, target both households with confirmed cases and those at elevated risk (e.g., sharing a water source). Additionally, these results warrant the use of vaccines to prevent both symptomatic and asymptomatic infections.

## Data Availability

All analytical code and de-identified data are available at github.com/UNCIDD/vc-household.

## Acknowledgements

This research was supported through programs funded by the National Institutes of Health, including the National Institute of Allergy and Infectious Diseases (AI168389 [K.E.W.], AI106878 [E.T.R. and F.Q.], AI099243 [J.B.H.], AI137164 [R.C.C. and J.B H.]), and Emerging Global Leader Award TW010362 (T.R.B.). The icddr,b is grateful to the Governments of Bangladesh and Canada for providing unrestricted/institutional support. The funders had no role in study design, data collection and analysis, decision to publish, or preparation of the manuscript. Data available at: github.com/UNCIDD/vc-household.

## Author contributions

Conceptualization: KEW, CPS, JL, ASA. Data curation: TRB, TI, FA, FC, AIK, RCL, RCC, AAW, SBC, ETR, JBH, FQ, KEW. Formal analysis: CPS, JL. Funding acquisition: KEW, SBC, ETR, FQ, JBH, RCC, TRB. Methodology: CPS, JL. Project administration: KEW. Software: CPS. Validation: CPS, JL, ASA, KEW. Visualization: CPS. Writing – original draft: CPS. Writing – review & editing: All authors.

