## Supplementary Materials for "Estimating the Transmission Potential of Symptomatic and Asymptomatic Cholera Cases from Household Microbiological and Clinical Data"

<u>Supplemental Methods</u>

*Model Specification*

The probability of an individual, *i*, residing in any given state, *n*, is calculated for every time step, *t*, and is denoted $\alpha_t^i(n)$. For enrolled household members, at a subset of time points culture results, vibriocidal titer, and/or symptom status are observed. Let $y_{i,t}^c$ , $y_{i,t}^v$ , and $y_{i,t}^s$ denote individuals *i*'s observed culture result (positive [$y_{i,t}^c = 1$] or negative [$y_{i,t}^c = 0$]), vibriocidal titer (≥320 [$y_{i,t}^v = 1$] or <320 [$y_{i,t}^v = 0$]), and symptom status (symptomatic [$y_{i,t}^s = 1$] or asymptomatic [$y_{i,t}^s = 0$]) at time *t*. For time steps where such an observation exists, the state probability is multiplied by an observation component representing the probability of the observation conditional on being in the given state.

The transition probability from state *m* to state *n*, $\tau_{mn}$, is stored in the transition matrix *T*, where the row gives the starting state and the column gives the ending state.

$$T = \begin{array}{c} \\ S \\ E \\ I_a \\ I_s \\ R \end{array} \begin{array}{c} \begin{array}{ccccc} S & E & I_a & I_s & R \end{array} \\ \left[\begin{array}{ccccc} 1 - f(I_a, I_s) - \frac{\epsilon}{4} & (1-\theta) f(I_a, I_s) - \frac{\epsilon}{4} & \theta(1-\phi) f(I_a, I_s) - \frac{\epsilon}{4} & \theta\phi f(I_a, I_s) - \frac{\epsilon}{4} & \epsilon \\ \epsilon & 1 - \sigma - \frac{2\epsilon}{3} & (1-\phi)\sigma - \frac{2\epsilon}{3} & \phi\sigma - \frac{2\epsilon}{3} & \epsilon \\ \epsilon & \epsilon & 1 - \gamma - \frac{3\epsilon}{2} & \epsilon & \gamma - \frac{3\epsilon}{2} \\ \epsilon & \epsilon & \epsilon & 1 - \gamma - \frac{3\epsilon}{2} & \gamma - \frac{3\epsilon}{2} \\ \epsilon & \epsilon & \epsilon & \epsilon & 1 - 4\epsilon \end{array}\right] \end{array} \quad (1)$$

For each individual *i* at time *t*, the function $f(I_a,I_s)$ gives the probability of becoming infected by at least one of the extra- and intra-household sources (Equation 2). We denote the extra-household infection risk for individual *i* as $b_i$, the intra-household infection risk from symptomatic individuals as $p_i^s$, and intra-household infection risk from asymptomatic individuals as $p_i^a$.

$$f(I_a, I_s) = 1 - (1 - b_i) \prod_{j \neq i} \left[\alpha_{t-1}^j(I_a)(1 - p_i^a) + \alpha_{t-1}^j(I_s)(1 - p_i^s) + \alpha_{t-1}^j(S) + \alpha_{t-1}^j(E) + \alpha_{t-1}^j(R)\right] \quad (2)$$

The intra- and extra-household infection probabilities are modeled as logistic functions of individual and household-level covariates, $X_i$ (Equations 3-5). Coefficients are shared between the intra-household probabilities for symptomatic and asymptomatic individuals, with the probability of infection for asymptomatic individuals containing an additional intercept term $\beta_{0,a}^p$ to account for the relative difference in infection probability of asymptomatic cases compared to symptomatic cases.

$$logit(b_i) = \beta_0^b + X_i\beta^b \tag{3}$$

$$logit(p_i^a) = \beta_0^p + \beta_{0,a}^p + X_i\beta^p \tag{4}$$

$$logit(p_i^s) = \beta_0^p + X_i\beta^p \tag{5}$$

The proportion of individuals with an incubation period less than one day is θ and is set at 30% in the main analyses based on estimates of the distribution of the incubation period [1]. For those individuals with incubation periods greater than one day, the rate of transition from the exposed to infectious compartment is σ. Once infected, an individual's probability of developing symptoms is $\phi$. Individuals transition from infectious to recovered compartments with probability $\gamma$. Once recovered, individuals are assumed to be immune to future infections for the duration of follow-up. All other transitions are assumed to occur with probability $\epsilon = 1 * 10^{-10}$.

The state probabilities $\alpha_t^i(n)$ are calculated using the forward algorithm:

$$\alpha_t^i(n) = \left[\sum_{m \in M} \alpha_{t-1}^i(k) * \tau_{mn}\right] * \omega_c(y_{i,t}^c, n) * \omega_c(y_{i,t}^v, n) * \omega_s(y_{i,t}^s, n) \tag{6}$$

Here $\omega_c(y_{i,t}^c, n)$ is the probability of the observed culture result for individual $i$ at time $t$ given the individual is in state $n$. Similarly, $\omega_v(y_{i,t}^v, n)$ and $\omega_s(y_{i,t}^s, n)$ give the conditional probability of the observed vibriocidal titer and symptom status, respectively. In the absence of an observation at time $t$, the corresponding observation component $\omega$ is set equal to 1. When an observation is available, the values of are taken from the observation probability matrix $\Omega_c$ for culture observations, $\Omega_v$ for vibriocidal titer observations, and $\Omega_s$ for symptom observations.

$$\Omega_c = \begin{array}{c} \\ y_{i,t}^v{=}0 \\ y_{i,t}^v{=}1 \end{array} \begin{array}{ccccc} S & E & I_a & I_s & R \\ s_- & s_- & 1-s_+ & 1-s_+ & s_- \\ 1-s_- & 1-s_- & s_+ & s_+ & 1-s_- \end{array} \tag{7}$$

Where $s_+ = 82.0\%$ is the sensitivity of culture and $s_- = 94.3\%$ is the specificity of culture [2].

$$\Omega_v = \begin{array}{c} \\ y_{i,t}^v{=}0 \\ y_{i,t}^v{=}1 \end{array} \begin{array}{ccccc} S & E & I_a & I_s & R \\ 1-\epsilon & 1-\epsilon & 0.9 & 0.9 & 0.3 \\ \epsilon & \epsilon & 0.1 & 0.1 & 0.7 \end{array} \tag{8}$$

$$\Omega_s = \begin{array}{c} \\ y_{i,t}^s{=}0 \\ y_{i,t}^s{=}1 \end{array} \begin{array}{ccccc} S & I_a & I_s & R & \\ 1-\psi & 1-\psi & 1-\epsilon & \epsilon & 1-\psi \\ \psi & \psi & \epsilon & 1-\epsilon & \psi \end{array}$$

(9)

Where $\psi$ is the probability of exhibiting cholera-like symptoms (watery diarrhea) when not infected. For the main analyses, we set $\psi$ = 13.0% based on the proportion of household contacts with neither a positive culture results or four-fold rise in vibriocidal titer who reported symptoms during the follow-up period.

We start the model one day before enrollment of the index household member to account for the fact that these individuals are already a portion of the way into the duration of their illness when they present for care. Index household members are assumed to start in state $I_s$ with probability 90%, state $S$ with probability θ *10%, and state E with probability (1-θ) *10%. The starting state probabilities for all other household members were fit.

The log likelihood is equal to the sum of the log state probabilities at the final time step ($t = 31$) across all enrolled and unenrolled members of participating households, $i = 1,2,...,N$, and states, $n \in \{S, E, I_a, I_s, R\}$.

$$L = \sum_{i=1}^{N} \sum_{n} \log[\alpha_{31}^{i}(n)] \tag{10}$$

Normally distributed priors with mean -3 and standard deviation 3 were used for logit-scale intercept terms and standard normal priors were used for logit-scale coefficients. Starting state probabilities for household contacts used a Dirichlet prior with all parameters set at 0.5. A normal prior with mean 0.9 and standard deviation 0.5 was used for logit-transformed σ and a normal prior with mean 0 and standard deviation 0.5 was used for logit-transformed γ.

*Model Validation*

We simulated ten independent datasets, each containing 500 households ranging in size from two to eight household members. In the underlying simulations, the daily probability of infection from a symptomatic household member was 5% and the and from an asymptomatic household member was 2.5%, implying a relative odds of infection of 0.49 for asymptomatic compared to symptomatic household members. The daily probability of extra-household infection was set at 1%, the proportion of infections that develop symptoms at 40%, the probability of having symptoms if not infected at 17%, the proportion of individual with a latent period less than one day at 30%, the duration individuals remain in the exposed compartment at 1.5 days, and the duration of infectiousness at 2 days.

For each simulated dataset and parameter, we calculated a 95% credible interval from the posterior distribution. Model accuracy was assessed via credible interval coverage – the proportion of credible intervals for a given parameter that contained the true value.

Additionally, we compared the number of people in each state at each time point in the underlying simulated data to the number of individuals implied to be in each latent state by the model, which was calculated by summing the state probabilities $\alpha_i^t(n)$ across all individuals, $i$, for each state $n \in \{S, E, I_a, I_s, R\}$ at each time point $t$.

Supplemental Tables and Figures

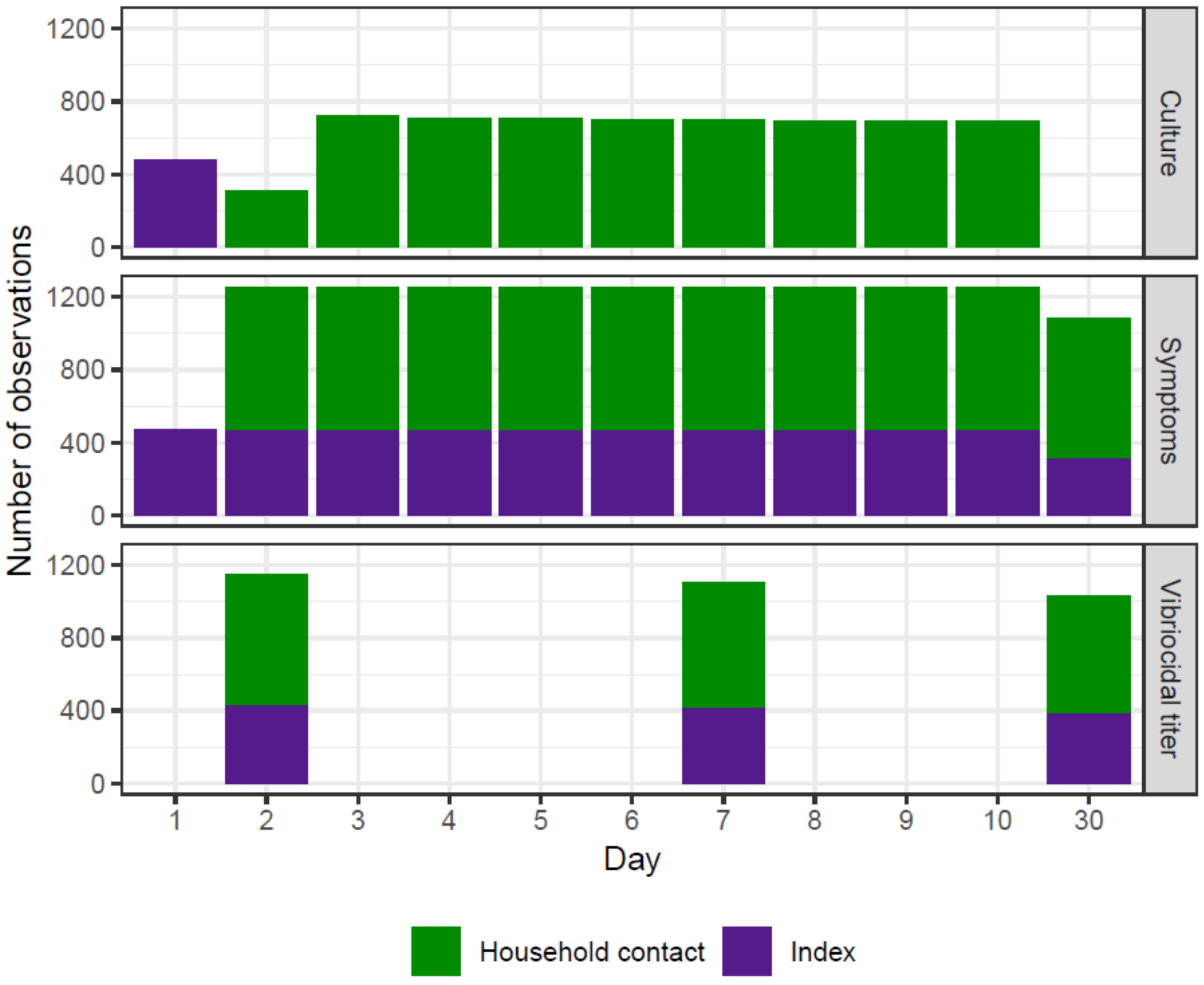

Figure S1: Number of observations on each sampling day for index cases and household contacts.

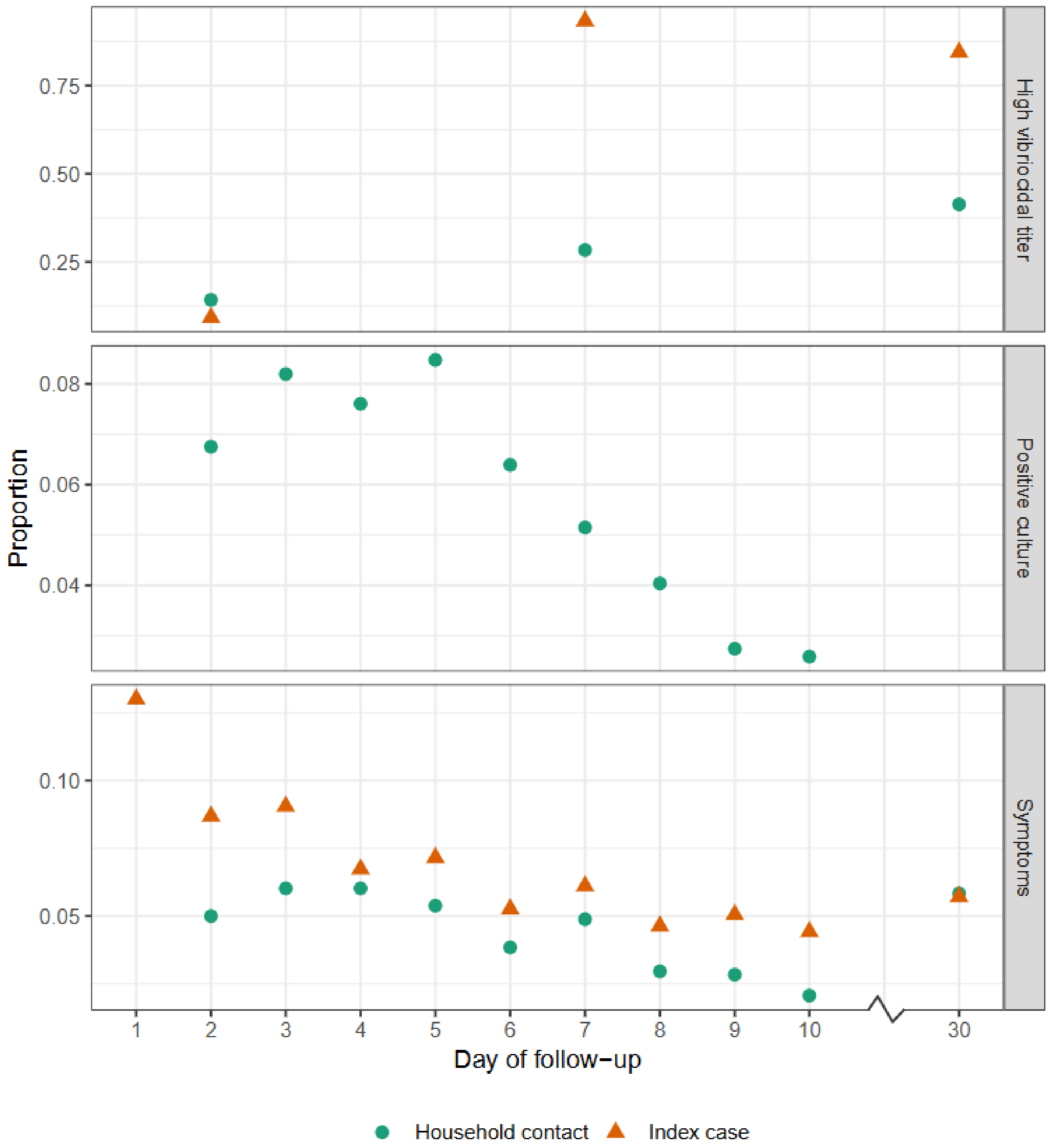

Figure S2: Proportion of index cases and household contacts with high vibriocidal titer (≥320), positive culture result, and symptoms over time.

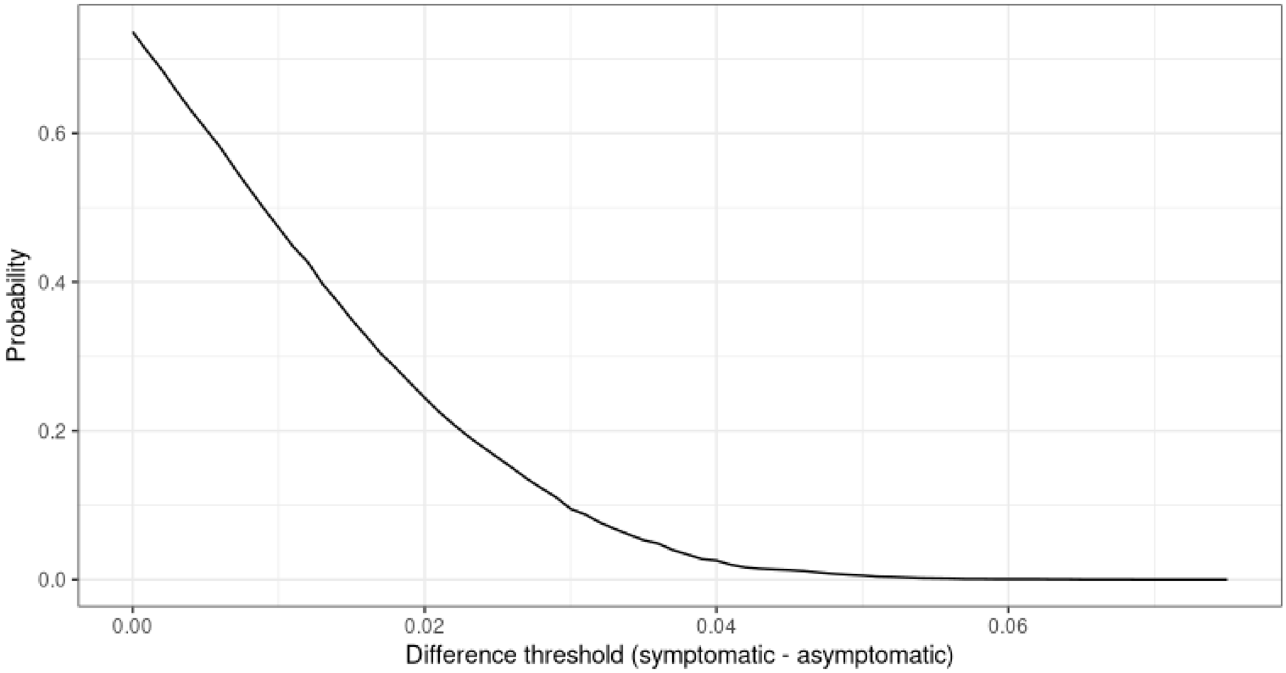

Figure S3: Probability that the per day probability of infection from a symptomatic household member is more than a given threshold greater than the per day probability of infection from an asymptomatic household member based on the posterior distribution of the baseline model.

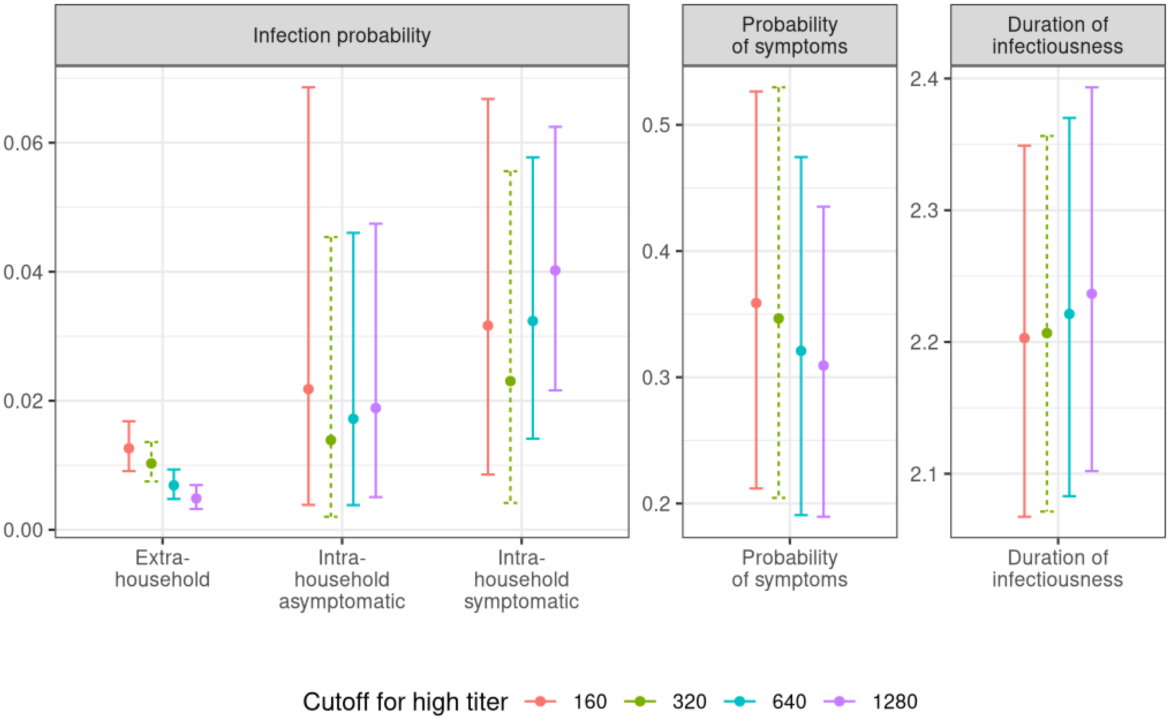

Figure S4: Sensitivity of daily infection probabilities, symptomatic proportion, and the duration of infectiousness to varying cutoffs for a high vibriocidal titer observation. The dashed line indicates the value used in the main analysis.

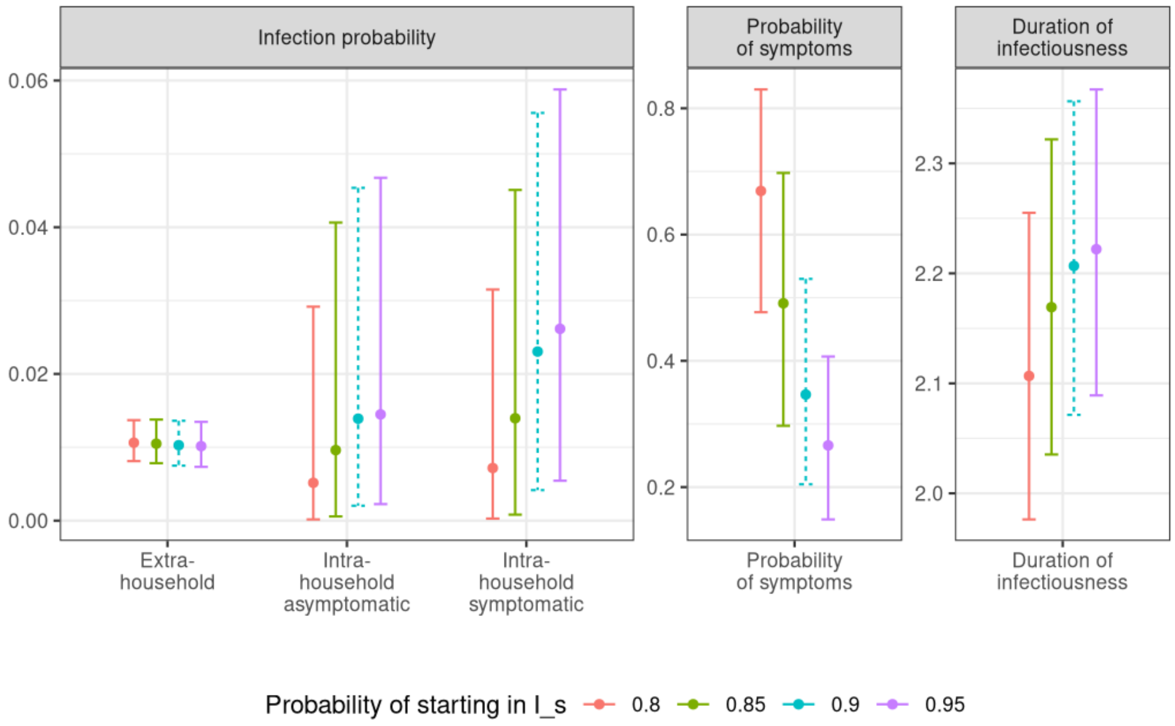

Figure S5: Sensitivity of daily infection probabilities, symptomatic proportion, and the duration of infectiousness to the probability that index cases start in the symptomatically infected compartment on day one of model time (which corresponds to the day before enrollment). The remaining probability is allocated to starting in the susceptible and exposed compartments, with the allocation between the two determined by the proportion of incubation periods that are less than one day (θ). The dashed line indicates the value used in the main analysis.

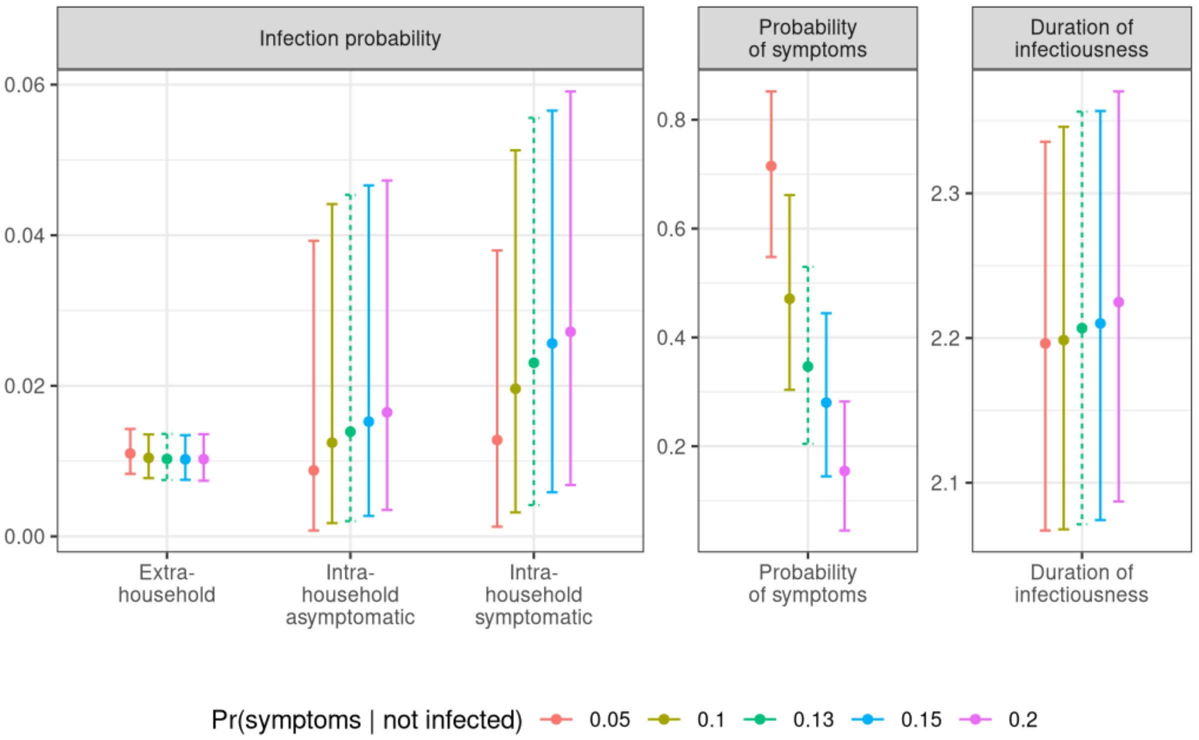

Figure S6: Sensitivity of daily infection probabilities, symptomatic proportion, and the duration of infectiousness to the probability of observing symptoms in uninfected individuals (ψ). The dashed line indicates the value used in the main analysis.

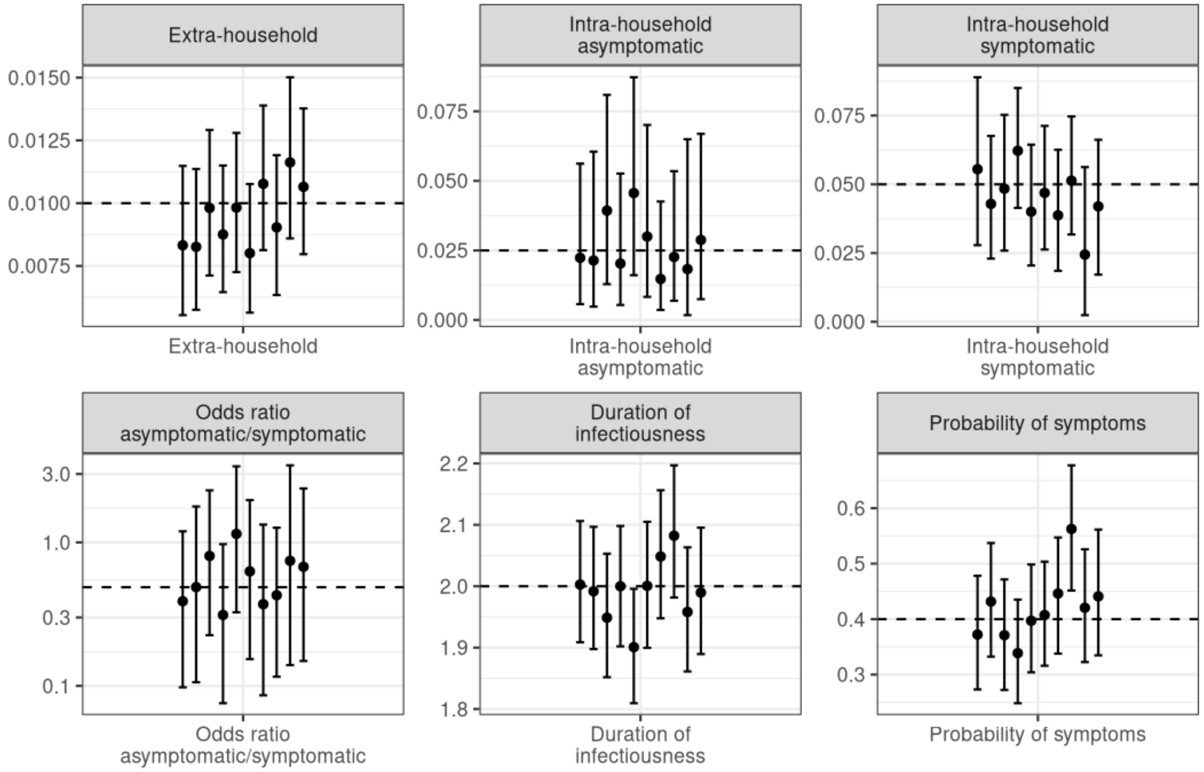

Figure S7: Parameter estimates from 10 model validation runs on simulated data. The dashed horizontal line shows the true value of each parameter.

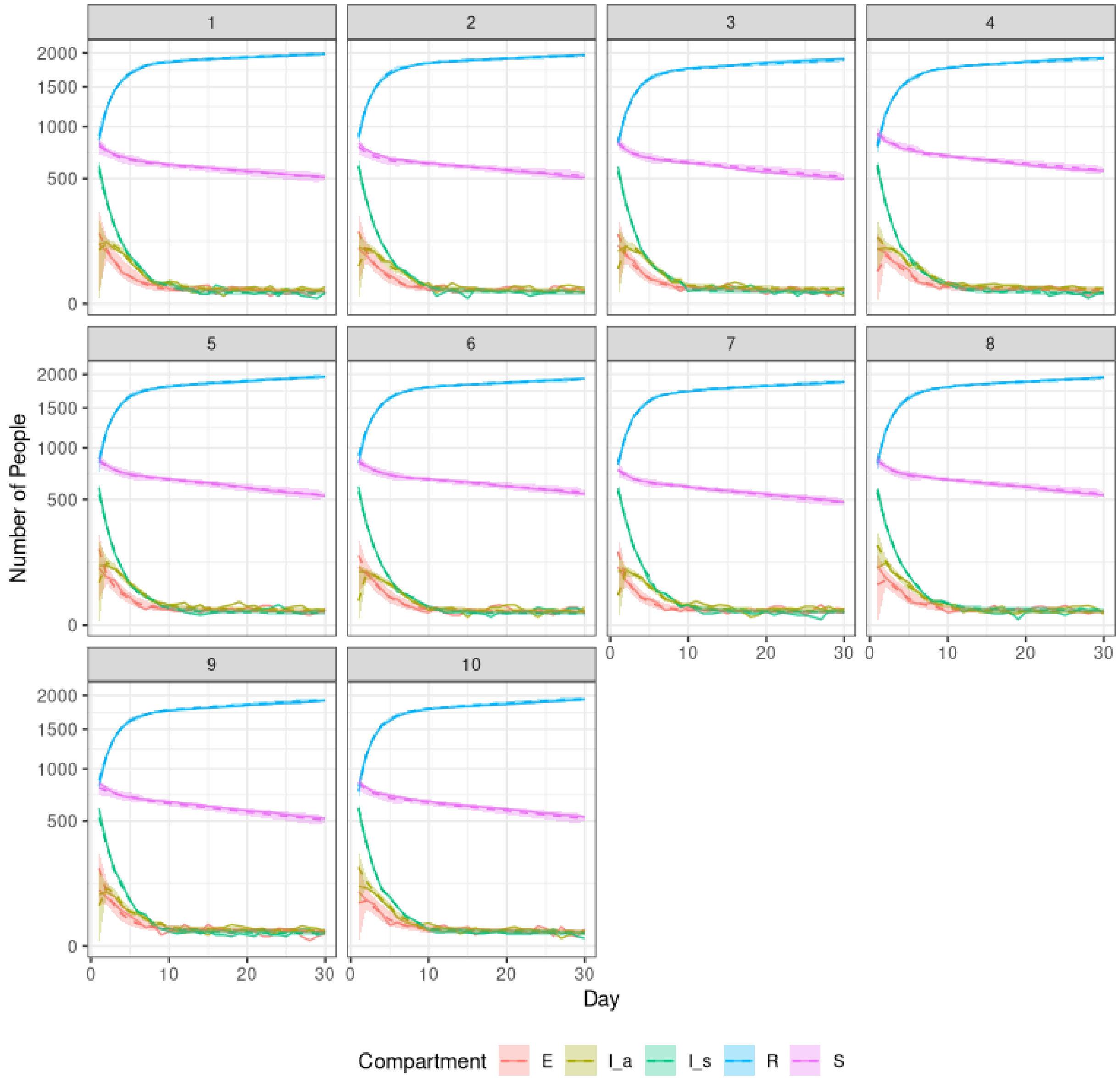

Figure S8: True number of people in each state (solid line) and number of people in each compartment implied by model (dashed line) and 95% credible interval for 10 model validation runs on simulated data.

| | Adjusted | | Unadjusted | |
|---|---|---|---|---|
| | Odds ratio (95% CI) | | Odds ratio (95% CI) | |
| | Intra-household | Extra-household | Intra-household | Extra-household |
| Sex | | | | |
| Male | Ref | Ref | Ref | Ref |
| Female | 0.99 (0.22-4.59) | 1.03 (0.52-2.08) | 1.15 (0.32, 3.98) | 1.33 (0.78-2.37) |
| Unknown | 1.01 (0.14-7.49) | 1.01 (0.15-6.79) | 0.80 (0.13, 5.13) | 0.71 (0.12-4.11) |
| Drinking water source | | | | |
| Boiled water | Ref | Ref | Ref | Ref |
| Private tap | 1.92 (0.38-7.55) | 1.62 (0.85-3.15) | 2.17 (0.51, 7.86) | 1.38 (0.73-2.66) |
| Tubewell | 1.24 (0.21-5.86) | 1.44 (0.75-2.89) | 1.58 (0.26, 6.03) | 1.69 (0.87-3.19) |
| Public tab or other | 0.78 (0.13-4.17) | 0.51 (0.11-1.93) | 0.73 (0.11, 3.54) | 0.49 (0.11-1.69) |
| Access to soap in home | | | | |
| Yes | Ref | Ref | Ref | Ref |
| No | 0.65 (0.12-3.05) | 2.03 (0.89-4.41) | 0.70 (0.13, 2.65) | 2.08 (0.86-4.27) |
| Monthly household income (BDT) | | | | |
| 0-8,499 | Ref | Ref | Ref | Ref |
| 8,500-11,999 | 0.90 (0.17-4.05) | 0.87 (0.45-1.59) | 0.78 (0.19-2.99) | 0.89 (0.46-1.70) |
| 12,000-17,999 | 0.86 (0.19-4.28) | 0.56 (0.26-1.06) | 0.86 (0.24-4.02) | 0.54 (0.25-1.10) |
| 18,000+ | 0.48 (0.10-2.40) | 0.42 (0.20-0.83) | 0.38 (0.10-1.57) | 0.45 (0.22-0.91) |
| Occupation group | | | | |
| Work outside home | Ref | Ref | Ref | Ref |
| Work at home | 0.77 (0.13-3.49) | 2.33 (1.13-5.31) | 0.79 (0.14-2.73) | 1.91 (1.06-3.37) |
| Under 18 | 1.31 (0.24-7.09) | 1.50 (0.57-4.12) | 2.41 (0.73-7.31) | 1.50 (0.73-2.96) |
| Unknown | 1.00 (0.14-7.76) | 0.96 (0.14-7.37) | 0.89 (0.13-5.54) | 0.81 (0.13-4.89) |
| Relation to index | | | | |
| Parent | Ref | Ref | Ref | Ref |
| Spouse | 1.03 (0.18-4.46) | 0.65 (0.32-1.22) | 1.10 (0.21, 3.78) | 0.71 (0.33-1.34) |
| Child | 1.84 (0.34-8.29) | 0.77 (0.25-2.14) | 2.28 (0.56, 7.27) | 0.82 (0.35-1.66) |
| Sibling | 1.33 (0.23-5.70) | 1.50 (0.59-3.36) | 1.87 (0.29, 6.64) | 1.32 (0.54-2.61) |
| Unknown or self | 1.17 (0.16-7.60) | 1.50 (0.30-5.27) | 1.24 (0.17, 6.64) | 1.09 (0.25-3.36) |
| Age | | | | |
| 0-4 | | | 2.40 (0.25-11.22) | 2.12 (0.37-8.32) |
| 5-17 | | | 1.91 (0.51-5.38) | 1.14 (0.55-2.05) |
| 18+ | | | Ref | Ref |
| Unknown | | | 0.92 (0.13-5.89) | 0.76 (0.12-4.60) |

Table S1: Adjusted and unadjusted odds ratio for intra- and extra-household infection risk by participant and household characteristic. The adjusted model includes all listed covariates except age.
